# Idiopathic subglottic stenosis arises at the interface of host and pathogen

**DOI:** 10.1101/2022.02.02.22270308

**Authors:** Alexander Gelbard, Meghan H. Shilts, Britton Strickland, Kevin Motz, Hsiu-Wen Tsai, Helen Boone, Wonder P. Drake, Celestine Wanjalla, Paula Marincola Smith, Hunter Brown, Marisol Ramierez, James B. Atkinson, Jason Powell, A John Simpson, Seesandra V. Rajagopala, Simon Mallal, Quanhu Sheng, Alexander T. Hillel, Suman R. Das

## Abstract

Idiopathic subglottic stenosis (iSGS) is a rare fibrotic disease of the proximal airway affecting adult Caucasian women nearly exclusively. Life-threatening ventilatory obstruction occurs secondary to pernicious subglottic mucosal scar. Diverse diseases in divergent organ systems are associated with fibrosis, suggesting common biologic mechanisms. One well characterized pathway is chronic inflammation secondary to pathogen. In the present study, we explored the role of the proximal airway microbiome in iSGS pathogenesis. In human samples, abundant bacteria are detectable in iSGS scar as well as in health subglottic controls or patients that developed subglottic stenosis following endotracheal intubation. Interestingly, the community structure of the iSGS proximal airway microbiome does not appear disrupted. Rather, in iSGS defects in the airway epithelial barrier allow displacement of the native microbiome into the immunoprivileged lamina propria and are associated with adaptive immune activation. Animal models of iSGS confirm both bacteria and an adaptive immune response are necessary for pathologic proximal airway fibrosis. Single cell RNA sequencing of the affected airway in iSGS offers an unbiased characterization of the observed epithelial barrier dysfunction. The airway scar in iSGS patients demonstrates basal cell depletion and epithelial acquisition of a mesenchymal phenotype. The epithelial alterations are associated with the observed microbiome displacement, dysregulated immune activation, and localized fibrosis. These results refine our understanding of iSGS and implicate shared pathogenic mechanisms with distal airway fibrotic diseases.

## INTRODUCTION

Idiopathic subglottic stenosis (iSGS) is a rare(*1*) but devastating fibroinflammatory airway disease that occurs almost exclusively in adult, Caucasian women(*2*). The disease is characterized by mucosal inflammation and localized fibrosis resulting in life-threatening blockage of the upper airway(*3*). Current treatments are limited by high recurrence rates, and the majority of iSGS patients require frequent procedural interventions following their initial diagnosis(*4*). Given the significant emotional, physical, and financial costs associated with recurrent airway obstruction(*5*), most research efforts have focused on procedural techniques to improve airway patency(*6*). However, highly focused scientific approaches to identify key elements of iSGS disease pathophysiology are essential to developing less invasive and more durable treatments. Histologically, iSGS cases show a pronounced immune infiltrate and mucosal fibrosis(*7*).

Diverse diseases in divergent organ systems are associated with fibrosis, suggesting common biologic triggers. One of the most well-characterized triggers is chronic inflammation secondary to pathogen(*8*). Investigations in alternate pulmonary pathologies have associated alterations in the airway microbiome with tissue remodeling in cystic fibrosis(*9*), and disease progression in pulmonary fibrosis(*10*). Recent data suggest iSGS may share these pathogenic mechanisms with lower airway fibrotic diseases. Chronic inflammation in airway scar(*11*) and bacteria in superficial mucosal swabs(*12*) provide preliminary support to the hypothesis that an aberrant immune response directed against bacterial species participate in the airway fibrosis in iSGS.

In the present study, we interrogated the relationship between microbial species and iSGS disease. We show abundant bacteria in iSGS scar with a community structure indistinguishable from controls. Interestingly, in iSGS the native microbiome appears displaced into the immunoprivileged lamina propria and is associated with significant immune infiltration. Unbiased molecular interrogation of iSGS airway scar with single cell RNA sequencing confirmed barrier dysfunction characterized by basal progenitor loss and residual epithelial acquisition of a mesenchymal phenotype. Animal models confirmed an essential role for both bacteria and an adaptive immune response to tissue remodeling after mucosal injury. Our data suggests that in iSGS the native microbiome is displaced across a dysfunctional epithelial barrier into the immunoprivileged lamina propria driving an adaptive immune response specific for native bacterial species. These results refine our understanding of iSGS, implicate shared pathogenic mechanisms with distal airway fibrotic diseases, and open new avenues for therapy.

## RESULTS

### Characterization of mucosal microbiome in iSGS

iSGS patients possess obstructive mucosal scar in the proximal airway distal to the vocal cords (**Figure 1A**). Study patients were diagnosed according to standard clinical criteria(*2*) and described in supplemental Table 1. We first quantified the number of 16S rRNA gene copies in deep tissue biopsies via qPCR (**Figure 1B**) and detected consistent signal in both iSGS patients and disease controls (patients that developed subglottic stenosis following prolonged intubation: iLTS). iLTS patients had a significantly higher bacterial load compared to iSGS samples (mean iSGS copy number: 520,000 vs iLTS: 1,370,000; P<0.0001). We then preformed 16S rRNA sequencing for insight into bacterial community structure. For further analysis, 37/50 (74%) iSGS and 18/27 (67%) iLTS samples were retained after implementing a cutoff of 500 high-quality 16S reads (2 standard deviations above the maximum number of reads in any of the negative controls).

**Figure 1.**
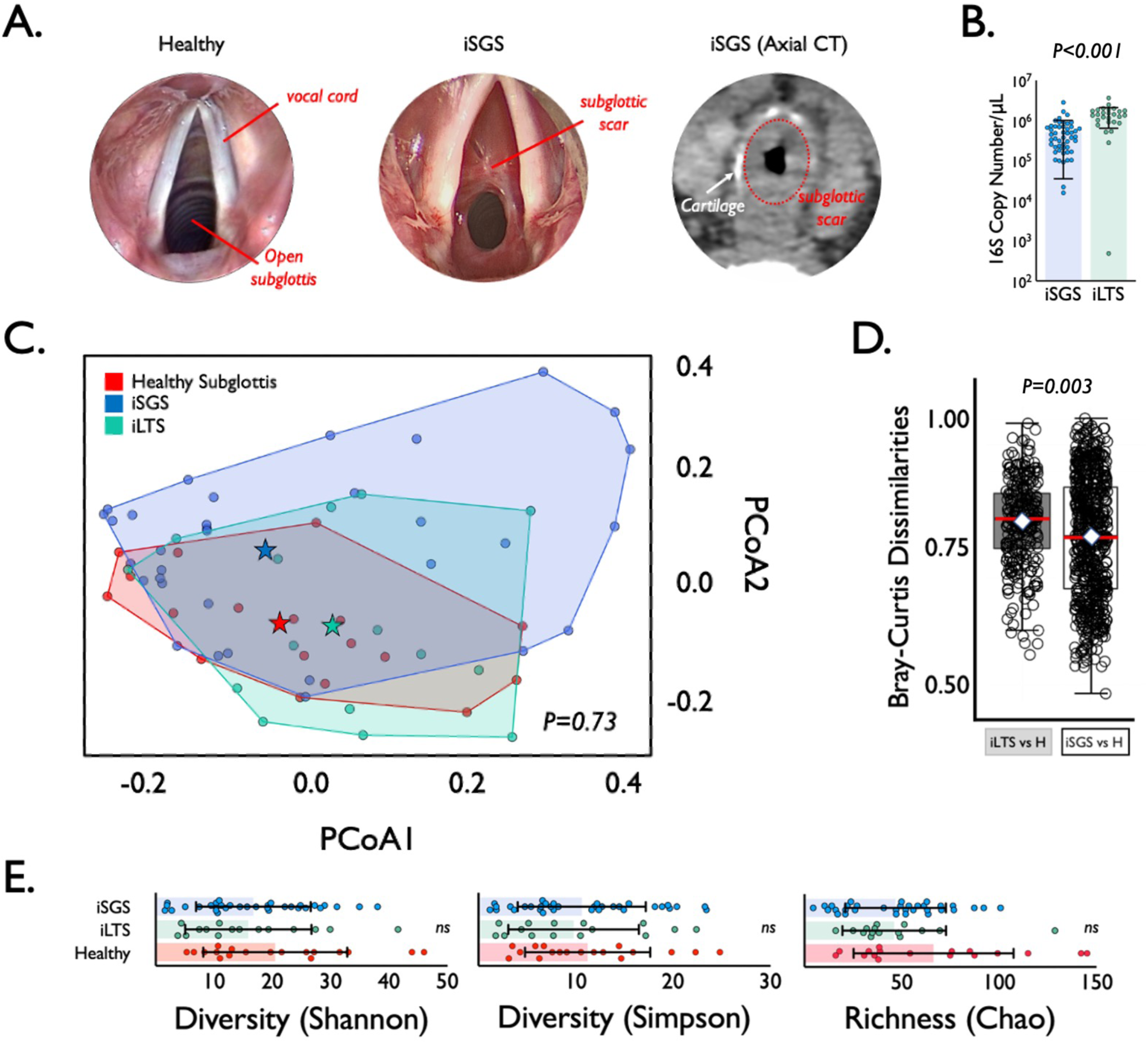
Microbial Speices in iSGS airway Scar. (A) Anatomy of mucosal scar obstructing the subglottis. Submucosal thickening with preserved cartilage seen on axial computed tomography. (B) qPCR of 16S rRNA showeda significantly higher bacterial load (copy number per μL) in iLTS patients (green) compared to iSGS samples (blue) (P<0.0001). (C) Principal coordinate analysis of proximal microbiome in iSGS, post-intubation subglottic stenosis (iLTS), and healthy controls showing no significant differences in the centroids between the three groups (PerMANOVA adonis2 testing p= 0.06). (D) Binary comparisons between healthy and iLTS samples, and between heathy and iSGS samples using Bray-Curtis dissimilarities showed iSGS samples more closely resembled healthy controls (Wilcoxon rank sum test with continuity correction, p-value=0.001). (E) Microbial alpha diversity (Shannon and Simpson indicies), and richness (Chao1 index) shown for iLTS, iSGS, and healthy control samples. Individual sample values are represented as dots. Group mean depicted by box center line, and standard deviation represented by error bars. Microbial alpha diversity and richness were not significantly different between cases and controls.

The top 20 most abundant bacterial families and genera are shown per each sample for iSGS, iLTS and healthy subglottic controls (**Supplemental Figure 1**). The majority of bacterial species present in healthy subglottis were consistent with the established healthy lung microbiome composed of supraglottic predominant taxa (e.g. *Prevotella*, *Streptococcus*)(*13*). Using principal coordinates analysis (PCoA), we compared the overall microbial community structure between iSGS, iLTS, and healthy subglottic controls. As seen in **Figure 1C**, there was no significant differences in the centroids between the three groups (PerMANOVA adonis2 testing p= 0.06). To validate these findings, we next utilized Bray-Curtis dissimilarities to make binary comparisons between only healthy and iLTS samples, and between only heathy and iSGS samples. iSGS samples more closely resembled healthy controls than iLTS samples resembled healthy subglottic controls (Wilcoxon rank sum test with continuity correction, p-value=0.001 **Figure 1D**). Additional testing of microbial community structure using established indices for diversity and richness confirmed no detectable differences between iSGS, iLTS and healthy controls. ANOVA testing of alpha diversity (mean <u>Shannon</u> index -iSGS: 15.28 vs iLTS: 14.53 vs healthy controls; p=0.39, mean <u>Simpson</u> index - iSGS: 15.28 vs iLTS: 14.53 vs healthy controls; p=0.822) and richness (mean <u>Chao1</u> index - iSGS: 40.75 vs iLTS: 40.45; vs. healthy controls, p=0.082, **Figure 1E**) showed no significant differences. Additionally, there was not a significant association between iSGS disease severity and overall bacterial load (p = 0.36, **Supplemental Figure 2A**) nor between disease severity and alpha diversity (p = 0.453, **Supplemental Figure 2B**) or richness (p-value = 0.6078, data not shown).

### Anatomic location of bacterial species in iSGS mucosal scar

To validate our findings and explore if superficial and deep tissue sampling methods produced unique bacterial communities, we next compared published 16S rRNA sequencing data of superficial swabs of iSGS scar (n=5)(*12*) with our deep tissue biopsies (**Figure 2A**). The top 20 most abundant genera were highly concordant between the superficial and deep sampling methods, despite differences in patient populations and lab processing protocols; offering support for our findings (the one significantly different genus was *Halomonas* which was abundant is swab samples and absent in tissue).

**Figure 2.**
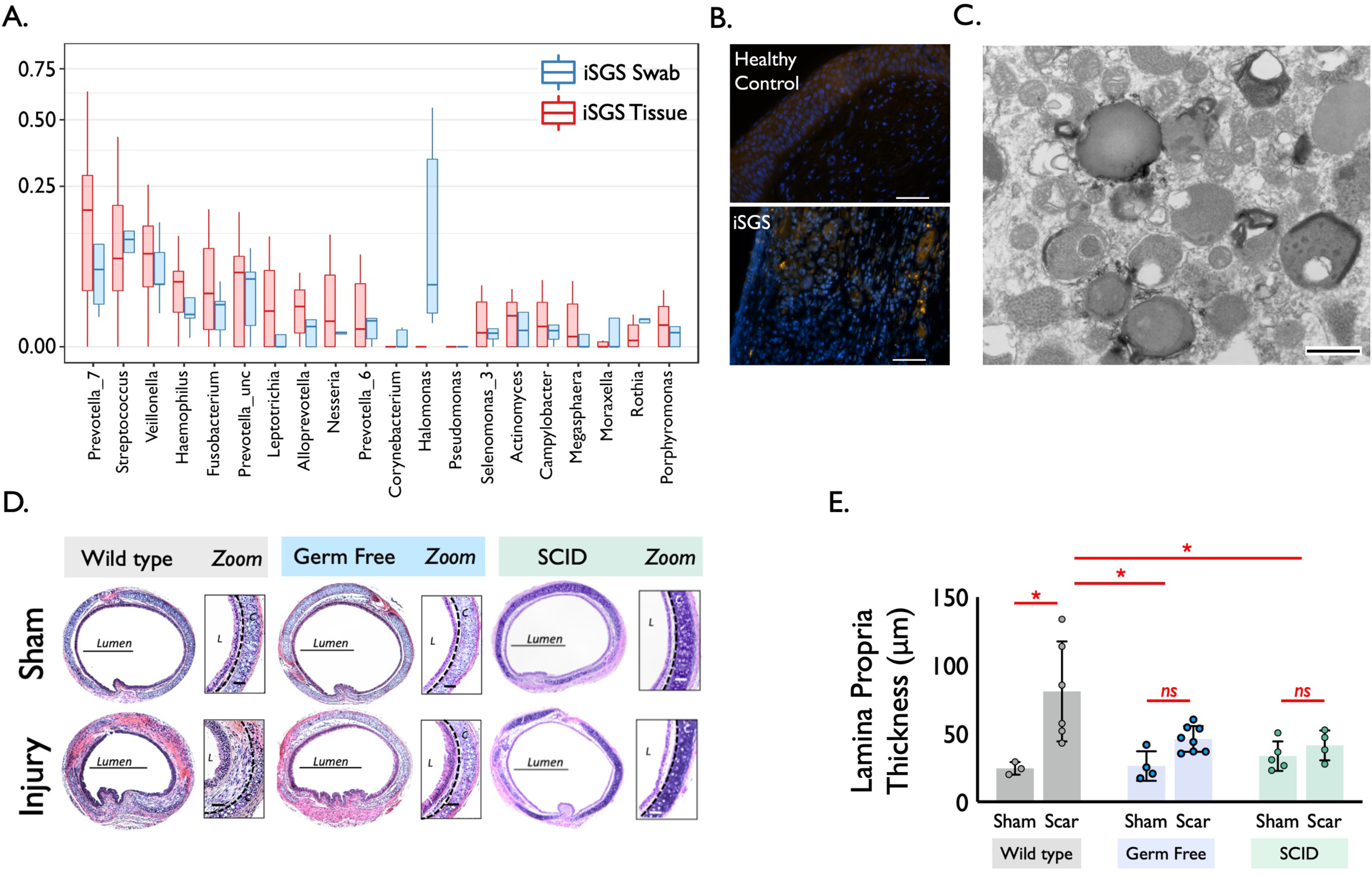
Requirement of both bacteria and host adaptivie immunity to generate airway scar following epithelial injury. (A) Comparison of bacterial abundance between sampling via mucosal swab vs. tissue biopsy in iSGS patients. The top 20 most abundant genera detected via the two methods with the genera abundance represented by boxplots; the median is represented by the center line of the box, and the interquartile range is represented by the upper and lower edges of the box. The vertical lines represent the whiskers (minimum and maximum, excluding outliers); outliers are represented as dots. Blue boxes are samples from Hillel et al., where the iSGS microbiome was sampled using swabs (N=5). Red boxes are samples from the tissue samples (N=37). Of the top 20 most abundant genera, the only significantly different genus between the two studies was *Halomonas*, which was relatively abundant in the iSGS swab samples but was not found in the iSGS tissue samples. (B) Fluorescence in situ hybridization with the pan-bacterial probe Eub338 shows iSGS mucosa possessed signal for bacteria in the deeper lamina propria while healthy control did not. Scale bar represents 50µm. (C) Transmission electron microscopy of separate iSGS scar specimen demonstrated numerous forms consistent with the size and shape of bacteria in the cell cytoplasm. Scale bar represents 500nm. (D) An established murine model of subglottic stenosis demonstrated significant thickening of the lamina propria 14 days after injury in wild type (WT) mice (p=0.0036). No significant thickening of the lamina propria was observed in either germ free mice (p=0.56), or severe combined immunodeficient mice (SCID) (p=0.98). (E) WT injury was significantly greater than both GF injury (p=0.025) and SCID injury (p=0.036).

Because 16S sequencing of the superficial and deep microbiome appeared identical, we investigated if native bacterial displacement into the deeper lamina propria was a unique feature of iSGS. Employing fluorescence in situ hybridization (FISH) with the pan-bacterial probe, Eub338, we investigated if mucosal biopsies from iSGS and healthy controls evidenced bacteria in the deep layers of the proximal airway mucosa. Representative FISH stains show iSGS mucosa possessed signal for bacteria in the deeper lamina propria while healthy control did not (**Figure 2B**). In a separate biopsy derived from iSGS mucosal scar, transmission electron microscopy demonstrated numerous forms consistent with the size and shape of bacteria in the cell cytoplasm, supporting the FISH staining (**Figure 2C**).

### Both bacteria and adaptive immunity are necessary for mucosal scar after epithelial injury

We next employed an established murine model of subglottic stenosis(*14*) to investigate the roles of bacteria and adaptive immunity in the mucosal fibrosis that characterizes iSGS. As expected, wild type mice developed mucosal inflammation and significant thickening of the lamina propria 14 days after injury (WT injury vs WT sham: 81µm +/- 37 vs. 24µm +/- 4, p=0.0036). Interestingly however, no significant thickening of the lamina propria was observed when using either germ free mice (GF injury vs GF sham: 46µm +/- 9.4 vs. 27 +/- 11, p=0.56), or severe combined immunodeficient mice (SCID)(*15*) lacking an adaptive immune response (SCID injury vs SCID sham: 41µm +/- 11 vs. 34 +/- 11, p=0.98). WT injury was significantly greater than both GF injury (p=0.025) and SCID injury (p=0.036) (**Figures 2D** **& E**).

### Single cell sequencing of subglottic mucosal scar in iSGS reveals epithelial cell loss and a pronounced immune cell infiltrate compared to unaffected mucosa

To determine the distribution and phenotype of the cellular populations present in iSGS airway scar, we generated single-cell suspensions from tissue biopsies of both airway scar (n=7) and matched unaffected airway mucosa (n=3) (Supplemental Table T2) and performed scRNAseq using the 10x Genomics Chromium platform (see supplementary Materials and Methods). The samples were collected and processed at two different sites (Supplemental Table T3); however, both sites collected cases and controls. In an effort to maximize our ability to identify rare cell populations, we jointly analyzed data from all samples. We defined inclusion criteria for cells based on observations from the entire dataset, removed low-quality cells accordingly, applied normalization and variance stabilization of the 25,974 recovered cells using Seurat(*16, 17*), integrated the data using the harmony(*18*), performed unsupervised clustering using Seurat, and classified the cell type of each cluster based on PanglaoDB(*19*) followed by manual annotation based on canonical markers to annotate clusters. We defined 22 cell types/states in the subglottis (initially one small CD8 effector T cell population was grouped together with a larger CD8 T effector population due to observations that cell cycle activity was driving distinct cluster identity). All cell types were identified both in airway scar and healthy mucosa. Notably, we did not observe overt batch effects driven by processing site or sequencing batch in our dimensionality reduction and visualization (Supplemental Figure S4). Cell types/states were also manually grouped into 4 broad tissue classes (Immune/Epithelial/Endothelial/Mesenchymal) based on their identity (**Figure 3A**), and confirmed with canonical lineage markers (**Supplemental Figure S5**). Quantification of cell types demonstrated significant differences between iSGS airway scar and matched healthy mucosa controls. Airway scar showed significantly more Immune cells (cell count per 1000 cells: scar vs healthy control: 636 vs. 238, P = 0.018) and significantly fewer epithelial cells (scar vs healthy control: 155 vs. 685, P < 0.001) (**Figure 3B**). The molecular results were confirmed at the protein level with flow cytometry (**Supplemental Figure S7**).

**Figure 3.**
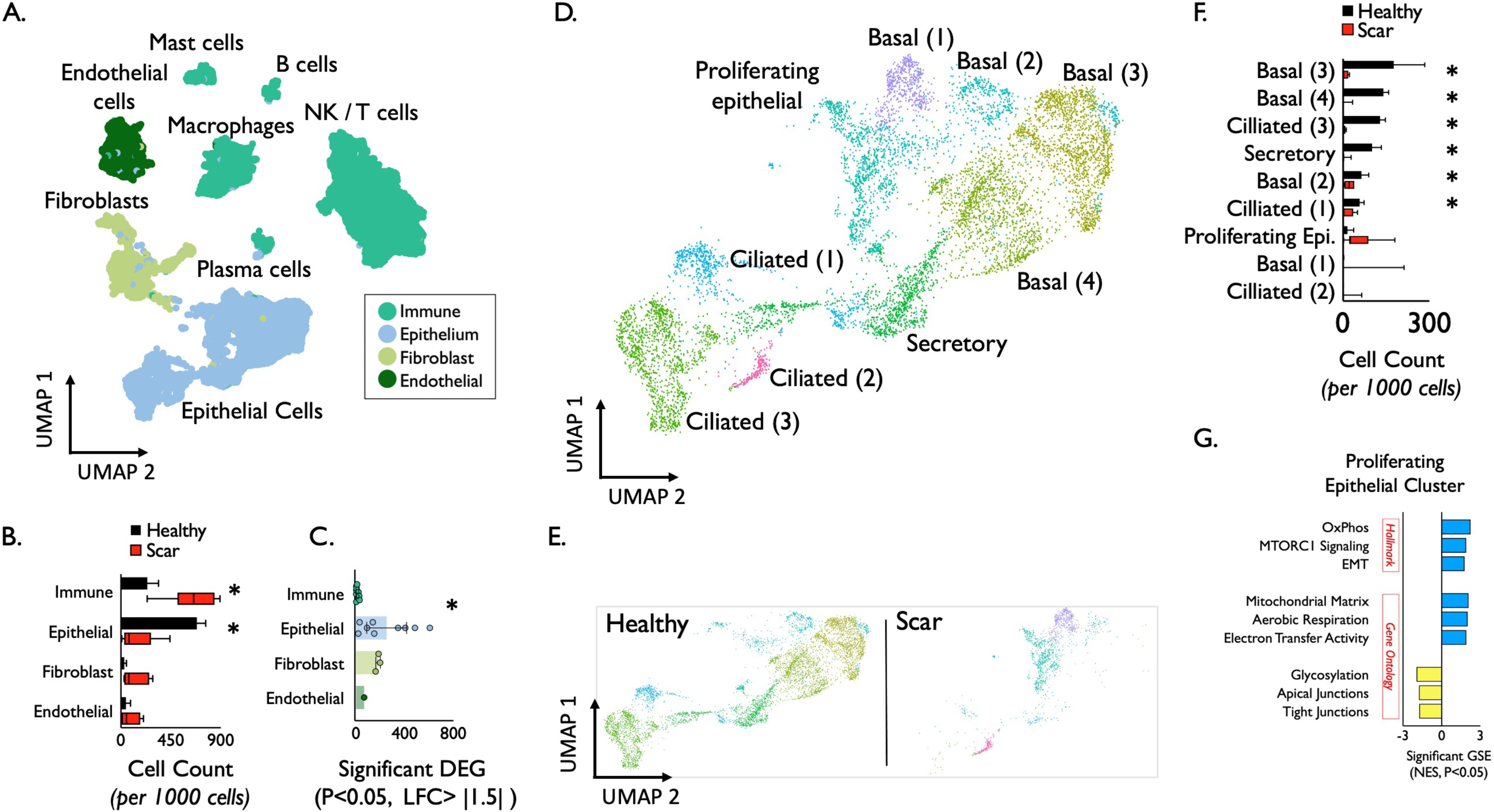
Epithelial cell identification and characterization in iSGS. Uniform Manifold Approximation and Projection (UMAP) of jointly analyzed single-cell transcriptomes from 25,974 cells from 7 iSGS mucosal scar and 3 healthy mucosa annotated by cell type. (A) Cell types/states manually grouped into 4 broad tissue classes. (B) Quantification of cell types showed significantly increased immune cell populations in airway scar (p = 0.018) and significantly reduced epithelial cell numbers (p < 0.001) Boxes depict median and interquartile range wiskers show min to max, *P < 0.05 by Mann-Whitney U. (C) Number of differentially expressed genes (DEG) in each cell type in iSGS airway scar and paired healthy mucosa [negative binomial test, log fold change (FC) cutoff of 1.5 and adjusted P value of <0.05]. (D) Detailed analysis of epithelial clusters identified conserved transcriptional programs in basal (four clusters), ciliated (three clusters), secretory (one cluster), and a proliferating cell subset (one cluster) in both healthy and scar (E). (F) Quantification of cell types in iSGS airway scar versus matched healthy mucosa. Boxes depict median and interquartile range wiskers show min to max, *P < 0.05 by Mann-Whitney U. (G) Differentially expressed gene (DEG) analysis for Hallmark genesets, Gene Ontology biological processes genesets in proliferating epithelial cells in iSGS airway scar compared to healthy mucosa (logFC, >0.5; FDR, <0.1). GSE: Geneset enrichment. Blue represents genesets upregulated in scar epitheluim, yellow represents genesets dwonregulated in scar epithelium compared to healthy mucosa. mTORC1: mechanistic target of rapamycin complex 1, OxPhos: oxidative phosphorylation, EMT: epithelial-mesenchymal transition.

In addition to quantitative differences in cell types/states, we compared phenotypic alterations between scar and healthy mucosa by examining the number of differentially expressed genes using EdgeR(*20*) (DEG: P<0.05, log fold change > |1.5|) (**Figure 3C**). This analysis demonstrated wide variability across the cell types, with epithelial cells and fibroblasts showing the greatest number of DEG (**Supplemental Figure S8**). When grouping cells into their tissue layer, the epithelium demonstrated significantly more DEG than immune cells (mean number of genes P < 0.05 and fold change > |1.5|, Epithelium = 252 vs 11 in immune, P = 0.007). The difference between epithelial cells and fibroblasts or endothelium was not significant. These results suggest that in addition to a quantitative reduction in cell numbers, the residual epithelium in iSGS scar is also phenotypically distinct.

### Molecular profiling of epithelium in iSGS scar revels significantly reduced basal populations and enrichment for a molecular program of epithelial mesenchymal transition

We further analyzed the epithelial clusters (**Figure 3D**) and identified conserved transcriptional programs in basal (four clusters), ciliated (three clusters), secretory (one cluster), and a proliferating cell subset (one cluster). Based on our observation of differential cluster abundance between scar and healthy mucosa (**Figure 3E**), we quantified the number of cell types/states from both scar and healthy mucosa. The clusters comprising basal, secretory, and ciliated cells showed significant reductions in scar samples (Boxes depict median and interquartile range, whiskers show min to max, *P < 0.05 by Mann-Whitney U.; **Figure 3F**). In addition to the dramatic loss of basal cells within airway scar, geneset enrichment analysis demonstrated that residual proliferating epithelial cells expressed a molecular program for epithelial mesenchymal transition (EMT) (**Figure 3G**). Additional upregulated genesets included oxidative phosphorylation and mTOR signaling consistent with observed proliferation markers Protein Phosphatase 1, Regulatory Subunit 105 (Ki67) and Cyclin Dependent Kinase 1 (CDK1) (**Supplemental Figure S5**). Gene ontology (GO) pathway analysis supported the Hallmark geneset EMT findings; iSGS airway scar showed enrichment for mitochondrial matrix genes (along with aerobic respiration and electron transfer activity). In parallel, proliferating epithelial cells in iSGS airway scar showed down-regulated glycosylation and junctional protein complexes (both apical and tight). Both aerobic respiration and loss of cell-cell adhesion are consistent with EMT.

### iSGS scar demonstrates increased adaptive immune cell subsets

Next, to explore the constituents of the immune cell infiltrate seen in iSGS airway scar (**Figure 4A**), we compared the cell types observed in scar with unaffected mucosa (**Figure 4B**). Quantification of cell types demonstrated significantly greater CD8 T_eff_ cells in scar (scar vs. healthy: 167+/-103 vs. 34+/26, P=0.033), along with more CD4+ T_reg_ (scar vs healthy: 37+/-15 vs. 4+/-3 P=0.005) and more NK cluster 2 cells (scar vs. healthy: 57+/-54 vs. 5+/-5, P=0.025). In contrast, the NK cluster 1 population was significantly reduced in scar (scar vs healthy: 4+/-9 vs. 36+/-37) (**Figure 4C**).

**Figure 4.**
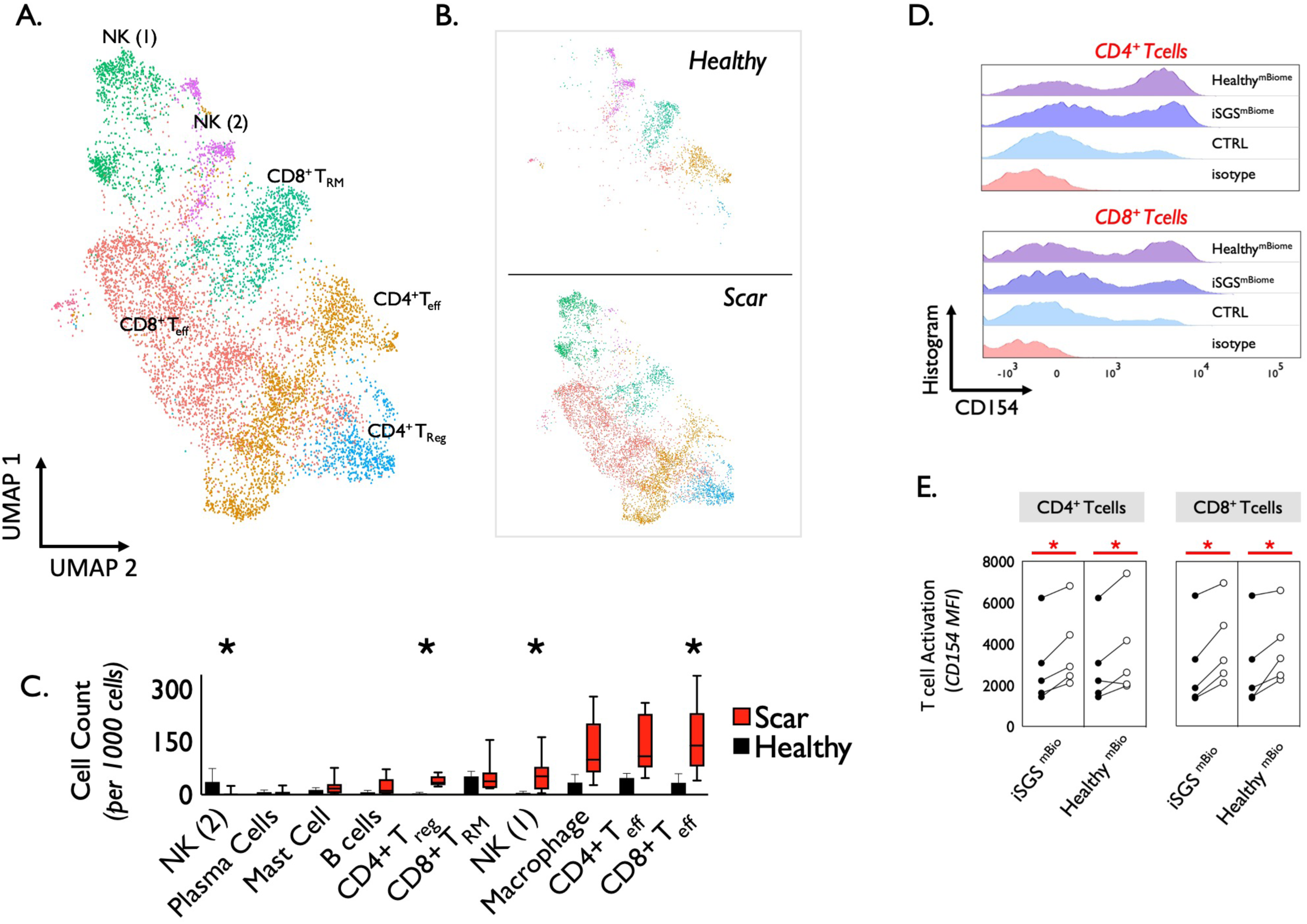
Immune cell identification and functional characterization in iSGS Proximal Airway Scar. (A) UMAP of jointly analyzed single-cell transcriptomes annotated by cell type displaying adaptive immune cells (T cells and NK cells). (B) Analysis of immune clusters identified conserved transcriptional programs in T cells (four clusters) and NK cells (two clusters), with differntial abundance in healthy mucosa and airway scar. (C) Quantification of immune cell types in iSGS airway scar versus matched healthy mucosa. Boxes depict median and interquartile range wiskers show min to max, *P < 0.05 by Mann-Whitney U. (D). Single cell suspensions from 5 unique iSGS patients cultured in the presence of matched iSGS airway microbiome, the microbiome from an unrelated healthy subject, or left untreated. 24 hours after stimulation, expression levels of activation marker CD154 were quantified on CD4+ and CD8+ T cells. (E) Both the matched iSGS microbiome, as well as the microbiome from an unrelated healthy control significantly up-regulated CD154 when compared to untreated experimental controls (CD4+ matched iSGS microbiome vs untreated: p=0.007; CD4+ unrelated healthy microbiome vs untreated: p=0.04), (CD8+ matched iSGS microbiome vs untreated: p=0.005; CD8+ unrelated healthy microbiome vs untreated: p=0.03). No significant difference observed between cells treated with a matched iSGS microbiome or cells treated with an unrelated healthy microbiome.

### The native proximal airway microbiome generates an antigen-specific immune response in infiltrating CD4+ and CD8+ T cells

In order to probe the function of the observed immune infiltrate, tissue biopsies acquired during operative endoscopy of 5 unique iSGS patients were used to create fresh single cell suspensions as described. Suspensions were rested for 6 hours, then cultured in the presence of a matched iSGS airway microbiome, the microbiome from an unrelated healthy subject, or left untreated. After 24 hours of stimulation, cells were washed and stained for markers of T cell activation (CD154) and analyzed via flow cytometry (**Figure 4D**). For CD4+ and CD8+ T cells, both the matched iSGS microbiome, as well as the microbiome from an unrelated healthy control significantly upregulated CD154 when compared to untreated experimental controls (CD4+ matched iSGS microbiome vs untreated: 3741 +/-1934, vs. 2912 +/- 1958, p=0.007; CD4+ unrelated healthy microbiome vs untreated: 3641+/- 2287 vs. 2912 +/- 1958, p=0.04), (CD8+ matched iSGS microbiome vs untreated: 3943 +/- 1989 vs. 2845 +/- 2098, p=0.005; CD8+ unrelated healthy microbiome vs untreated: 3784 +/- 1768 vs. 2845 +/- 2098, p=0.03) (**Figure 4E**). For both CD4+ and CD8+ T cells, CD154 expression was not significantly different between cells treated with a matched iSGS microbiome or cells treated with an unrelated healthy microbiome. These data suggest that while the native airway microbiome can trigger an antigen-specific immune response in iSGS, T cell activation in iSGS mucosal scar can also occur when presented with the bacterial constituents of a healthy individual.

## DISSCUSSION

Idiopathic subglottic stenosis (iSGS) is a debilitating localized fibrosis of the proximal airway. Affected patients possess tightly conserved clinical demographics, histopathology, and physiologic impairment(*4*). Our data suggest that defects in epithelial barrier function can allow displacement of the native microbial community deep into the airway mucosa and contribute to dysregulated immune activation leading to fibrotic remodeling in iSGS. Our animal models reveal that both bacteria and an intact adaptive immune response are necessary to drive tissue remodeling after proximal airway epithelial injury.

The pseudostratified epithelium lining the human airway is comprised of a number of distinct populations of cells with specialized effector functions. Airway epithelium and the overlying mucocilliary layer maintain a physical barrier against environmental insults (pathogens, allergens, and toxins). Many primary respiratory diseases, including chronic obstructive pulmonary disease (COPD), asthma, and idiopathic pulmonary fibrosis (IPF) display substantial pathological alterations in the airway epithelium. Evidence suggests impairment of the epithelial immune barrier allows bacteria to penetrate the airway surface liquid, intercalate within the epithelium, and activate host immunity. This inflammation, when sustained inappropriately, can culminate in fibrotic tissue remodeling and physiologic impairment.

In COPD secondary to cigarette exposure, respiratory mucosal inflammation and fibrotic remodeling contribute to small airway obstruction and clinical symptoms(*21*). Yet even after smoking cessation, many patients with COPD have sustained inflammation and disease progression(*22*). Published data in COPD demonstrate this persistent epithelial dysfunction (even after cessation of the inciting toxic insult) results from impaired immunbarrier protection. Dysfunctional epithelium leads to bacterial invasion deep into the mucosa and fibrotic airway remodeling(*23*). Animal models suggest that endogenous bacteria orchestrate a persistent and pathologic adaptive immune response that drives tissue remodeling(*24*).

Similarly, asthma is characterized by airway inflammation and tissue remodeling in response to antigenic trigger (*25*). In asthmatics, the epithelial barrier loses differentiation, and junctional integrity is impaired. Epithelial dysfunction appears to be a central driver of atopy, as it frequently pre-dates the development of allergic symptoms(*26*). In IPF, animal models, *in vitro* human data and genetic evidence suggest that the airway epithelium plays a central role in disease susceptibility and initiation(*27*). It is hypothesized that repetitive, small injuries to vulnerable alveolar epithelial cells result in apoptosis and impaired re-epithelialization, while epithelial-mesenchymal interactions drive subsequent extracellular matrix expansion(*28*).

Beyond the central role airway epithelial cells play in human respiratory disease, numerous studies have shown a specific role for the basal cell subset in repair and regeneration of the respiratory system after epithelial injury(*29–31*). Fate-mapped mouse strains have shown that basal cells contribute to regeneration of the pulmonary parenchyma in response to both chemical(*32*) and viral injury(*33*). Murine models of chemical-induced epithelial injury have shown that severe mucosal destruction involving the basal cell layer is associated with an uncontrolled proliferation of the underlying stroma, resulting in an accumulation of fibroblasts and immune cells that subsequently obliterate the airway lumen(*34*). The reduction of basal cell subsets we observed in our data may partially explain the discorded tissue remodeling seen clinically in iSGS(*7, 35*).

Interestingly, in addition to the observed basal cell depletion in iSGS airway scar, geneset enrichment analysis (GSEA) of the residual epithelial cells showed mechanistic target of rapamycin (mTORC1) pathway activation and enhanced aerobic metabolism. mTOR is a master sensor that integrates environmental factors to regulate cell growth. In general, activation of mTOR stimulates proliferation, mitochondrial biogenesis and oxidative phosphorylation(*36*). The GESA findings were consistent with the observed KI67 and CDK1 expression (marking cellular proliferation) in the epithelium of iSGS airway scar. Additionally GSEA showed the residual epithelium in iSGS airway scar acquired a mesenchymal phenotype with epithelial-mesenchymal transition (EMT) pathway enrichment. EMT allows disassembly of cell-cell junctions, actin cytoskeleton reorganization and induction of contractile proteins as non-motile epithelial surfaces convert into individual, a motile mesenchymal phenotypic. These phenotypic alterations may result from localized pathogen-driven inflammation(*37*) or can result from mucosal injury secondary to physiologically relevant bile acid exposure(*38*). Integrating the basal cell depletion with GSEA findings supports the hypothesis that mucosal barrier dysfunction participates in iSGS disease pathogenesis.

Although an often overlooked anatomic subsite, the subglottis is uniquely enriched in antigen-presenting dendritic cells as well as T lymphocytes (*39, 40*). Additionally, it functions as a transition zone from the ciliated lining of the trachea to the squamous epithelium of the larynx. As a consequence, the subglottis has increased exposure to pathogens as the cilia-driven upward movement of the airway mucus layer temporarily stalls(*41*). While the inciting event for barrier dysfunction in iSGS is unclear, both genetic predisposition and environmental insults can impair epithelial function. Although the genetic foundations of iSGS remain obscure(*42*), many candidate susceptibility genes identified via GWAS for Asthma(*43*) and COPD(*44*) suggest a role for epithelial damage and adaptive immune activation in disease pathogenesis. Similarly, validated susceptibility genes for IPF(*45*) frequently impact epithelial integrity(*46*). Alternatively, allergens are capable of disrupting physical integrity of the barrier via their protease activity, while several respiratory viruses specifically target junctional proteins in the airway epithelium leading to barrier dysfunction (*26*) and adaptive immune activation(*47*).

Prior small case series examining environmental factors contributing to iSGS have implicated disruption of the proximal airway microbiome, one using pathogen-specific molecular approaches(*48*), and another employing 16S rRNA sequencing of mucosal swabs(*12*). While these studies implicated the presence of microbial species in iSGS, our new data from a larger, and more diverse patient cohort provides an unbiased characterization of the mucosal tissue microbiome. Our current results suggest that rather than microbiome disruption, in iSGS a “healthy microbiome” is displaced across a dysfunctional epithelium. This displacement is associated with adaptive immune infiltration and activation in response to bacterial species. This is supported by the finding that both a matched host microbiome, as well as the microbiome from a healthy control can drive adaptive immune activation in the proximal airway scar.

However, the displaced microbiome may not be the sole target of the adaptive immune response observed in iSGS mucosal scar. A feed-forward inflammatory loop may become established when peptides from microbial proteins share sufficient structural similarity with self-peptides and activate autoreactive T cells, termed “molecular mimicry”(*49*). Inflammation resulting from bacterial infection can also activate local antigen-presenting cells and enhance processing and presentation of self-antigens, referred to as “epitope spreading”. Sustained innate immune activation in response to pathogen specific molecular patterns (PAMPs) may also participate in the observe localized mucosal inflammation in iSGS and potentiate epithelial damage.

Although we acknowledge the limited ability to assign causality in pathologic studies involving human tissue, combining bacterial profiling along with histologic localization, animal models and *in vitro* techniques provided consistent support to our hypothesis. Despite the inherent limits involved in rare disease research, our translational approach offers new insights into disease pathogenesis. Additionally, deconstructing the proximal airway tissue into its component cells based on transcriptional data derived from single cell RNA sequencing provides an unbiased view into the cellular ecosystem of human idiopathic subglottic stenosis, helping us to characterize the phenotype, major cell types in airway scar without the biases that are typically introduced by pre-selection of markers. Complementing this molecular data, our functional studies support the concept that defects in epithelial barrier function allow translocation of the native microbial community deep into the airway mucosa and drive dysregulated immune activation leading to fibrotic remodeling in iSGS. Animal models reveal that both bacterial and an intact adaptive immune response are necessary to drive tissue remodeling after proximal airway epithelial injury.

Our findings dramatically shift our concept of iSGS disease pathogenesis and implicate shared pathogenic mechanisms with distal airway fibrotic diseases. Most critically, these results suggest that proven treatment approaches pioneered in pulmonary fibrosis may provide therapeutic benefit in iSGS and warrant rigorous future study.

## METHODS

Detailed experimental methods are included in the Supplemental Material.

The accession number for the 16S sequencing data reported in this paper Bioproject number PRJNA784956, and the accession number for the single cell RNA-sequencing data is GSE191128.

## STUDY APPROVAL

Patients in this study with subglottic stenosis provided written and informed consent to tissue collection under a Vanderbilt University Medical Center institutional review board (IRB)-approved protocol (no. 140429). Healthy subglottic controls were obtained under UK Research Ethics Committee (no. 14/SS/1015 & 15/WM/0349). Human experiments at Johns Hopkins University were performed under IRB-approved protocol (no. NA_00078310). For murine experiments, all mice were housed and bred at Johns Hopkins in accordance with the regulatory standards of the National Institutes of Health (NIH) and American Association of Laboratory Animal Care standards and were consistent with the Johns Hopkins Institution of Animal Care and Use Committee (no. MO18M124).

## AUTHOR CONTRIBUTIONS

B.S., H.B., H.B., J.P. and J.S. performed 16S sequencing experiments. S.D. S.R. and M.S. conducted microbiome analysis. A.H., H.T. and K.M. performed murine experiments. P.M. preformed FISH assay. A.G. and C.W. participated in preparation and analysis in vitro T cell activation assays. A.G., M.R. and Q.S. processed and analyzed RNA-seq data. A.G. designed the experiments and wrote the manuscript. W.D. and S.M. provided critical review of experimental design and preformed data analysis.

## Data Availability

All data produced in the present study are available upon reasonable request to the authors.

## ACKNOWLEDGEMENTS

This work was supported in part by the NIH/NHLBI grant no. R01HL146401 (A.G.). Patient-Centered Outcomes Research Institute award number: 1409-22214 (A.G.).

## EXTENSIVE METHODS

### Case definitions

We diagnosed <u>Idiopathic Subglottic Stenosis</u> (iSGS) according to standard clinical criteria.^1, 2^ In brief, patients had no history of endotracheal intubation or tracheotomy within 2 years of presentation. They had no history of significant laryngotracheal injury, thyroid or major anterior neck surgery, or neck irradiation. They had no caustic or thermal injuries to the laryngotracheal complex. They had no clinical history of vasculitis and negative titers antinuclear cytoplasmic antibody (ANCA). Anatomically, the airway stenosis involved the subglottis. (Supplementary Table S1).

### Control definitions

<u>Post intubation Subglottic Stenosis (iLTS)</u>: Patients who developed subglottic or tracheal stenosis within 2 years of intubation or following tracheostomy.^1, 2^

<u>Healthy Subglottis:</u> Healthy control subjects underwent elective direct laryngoscopy under general anesthesia for a range of upper aerodigestive symptoms but normal subglottic examination and had subglottic mucosal biopsies collected using a sheathed cytology brush (BC-202D-5010; Olympus)

### Study Procedures

#### Microbiome Assessment

We performed tissue sampling of the affected airway in the operating room in patients undergoing surgical treatment for their airway stenosis. Mucosal brush biopsies from healthy controls were also obtained under direct endoscopic vision in the operating room during elective laryngoscopy under general anesthesia. In patients with mucosal scar undergoing tissue biopsy, after surgical exposure of the subglottis during direct laryngoscopy, the airway mucosa was washed with 10cc of sterile saline. After rinsing the mucosa sterile upbiting cup forceps were used for mucosal biopsy. Without any break in sterility, a researcher received the mucosal explant in the operating room immediately after transfer from the surgical field. Tissue samples were placed in sterile RNAlater solution and stored at -80°C. Similarly, subglottic mucosal brush biopsies from healthy controls were directly placed in sterile RNAlater solution and stored at -80°C.

### DNA extraction, quantitative PCR and 16S rRNA gene sequencing

#### DNA Extraction

DNA was extracted with the DNeasy PowerSoil Kit (Qiagen). Mechanical lysis of bacterial cell walls was performed by shaking the samples on a TissueLyser II (Qiagen) for 20 minutes total. Due to the expected low bacterial biomass of tissue samples, a total of 10 negative controls were processed concurrently with the samples (8 of which were extraction negative controls and 2 of which were PCR negative controls).

#### Determination of bacterial load by qPCR

DNA input was standardized by equal volumes due to standardized sample size and multiplexed DNA extraction methods. Equal volumes of DNA were added to triplicate reactions containing universal 16S rRNA primers (UniF340 actcctacgggaggcagcagt, UniR514 attaccgcggctgctggc)^3^, BioRad iQ Supermix, and Invitrogen SYBR DNA stain following the manufacturer’s protocol. Each qPCR plate included a corresponding extraction negative and a no-template negative control. A serial dilution of standards containing known bacterial copy numbers specific to the primer pair were used as a standard curve as previously described. PCR reactions were run using a BioRad CFX96 Real-Time PCR Detection System with a 15s 95°C melting and 1 minute 54°C annealing step for 40 cycles. CT values were plotted against the standard curve to determine copy number. Figure generation and statistical values were generated using Prism version 8.

### 16S rRNA Library Construction

Dual-indexed universal primers appended with Illumina-compatible adapters were used to amplify the hypervariable V4 region of the bacterial 16S rRNA gene.^4^ The PCR mix for each library contained 12.5 µl of MyTaq Mix (Bioline), 0.75 µl DMSO, 1 µl of forward primer, 1 µl of reverse primer, 7 µl of extracted DNA, and PCR Certified water (Teknova) was added to achieve a final volume of 25.25 µl. DNA was denatured at 95°C for 2 min, and then 30 cycles of 95°C for 20 seconds, 55°C for 15 seconds, and 72°C were performed. Samples were then incubated at 72°C for 10 min, and samples were held at 4°C until removal from the thermocycler. Each sample was run on a 1% agarose gel to verify reaction success. Libraries were cleaned and normalized with the Invitrogen SequalPrep Kit. After normalization to 1-2 ng/µl, 10 µl of each sample were combined to create the sequencing pool. The pool was cleaned with 1X AMPure XP beads (Beckman Coulter, Brea, California). Libraries were sequenced on an Illumina MiSeq with 2×250 bp reads. A mock community control (ZYMOBiomics) and extraction and PCR negative controls were run concurrently along with the samples to assess data quality and levels of background contamination.

#### 16S Sequence Processing

We processed the 16S rRNA sequences with the dada2 pipeline by following the standard operating procedure (available at: https://benjjneb.github.io/dada2/tutorial.html, as of 7.16.2019).^5^ Sequences were grouped into amplicon sequence variants (ASVs)^6^, and taxonomy was assigned using the SILVA reference database.^7^ Sequences were subsequently processed through the R package decontam^8^ to remove any suspected contaminants that were found in the negative control samples. Potential contaminants were detected with the decontam “prevalence” method, in which presence/absence of sequences in negative controls is compared to that of real samples. The R package phyloseq^9^ was used to facilitate downstream data processing. Figures were generated with the R package ggplot2.^10^

#### Comparison of tissue-based and swab-based sampling of microbial species in iSGS

In Hillel et al., superficial swabs of airway mucus were used to characterize the microbiome ^11^, while in our study we sampled the tissue directly. We downloaded the FASTQ files available from the first study to compare the superficial microbiome with the deeper tissue microbiome from in our study as described above.

### Fluorescence in situ Hybridization

Fluorescence in situ Hybridization (FISH) was performed as previously described^12^. In brief, custom oligonucleotides were generated by the Vanderbilt University Molecular Cell Biology Resource Core in partnership with Sigma/Genosys (Woodlands, TX). The pan-bacterial probe (Eub338) and negative control probe (Non-Eub) sequences are listed in Table 1, below. Three diseased tracheal specimens and one healthy control tracheal specimen in FFPE were used for this analysis. Human specimens were stained with custom oligonucleotides as previously described. FFPE blocks were cut into 5-micron sections on charged slides and placed in a hybridization oven at 50° C for 10 minutes to melt paraffin and then de-paraffinized in Histoclear^®^ (National Diagnostics) and re-hydrated by ethanol gradient before being placed in 20 mM Tris buffer. Bacterial probes were diluted to 2 μM in pre-warmed hybridization buffer (20 mM Tris-HCl [pH 8.0] + 0.9 M NaCl + 0.01% sodium dodecyl sulfate). Approximately 150 μL probe solution was then placed on each slide until sample was completely covered and slides were incubated for 1.5 hours at 46° C in humidity chamber. Slides were then washed in FISH wash buffer (225 mM NaCl + 20 mM Tris + 5 mM EDTA) for 5 minutes, three times, before being counterstained with DAPI (1:1,000 diluted in PBS) for 5 minutes. Slides were then washed again in FISH wash buffer for five minutes, twice. Slides were then cover slipped with ProLong Gold Antifade (Invitrogen) and allowed to cure in the dark for 24 hours prior to sealing.

**Table 1:**
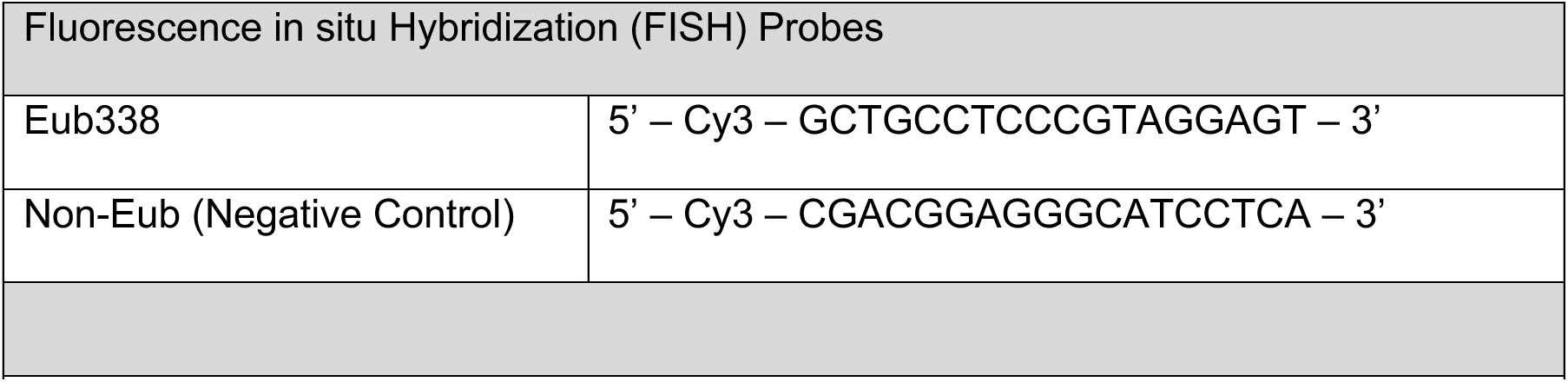
Fluorescence in situ Hybridization (FISH) probes utilized.

### Transmission Electron Microscopy (TEM)

Specimens were processed for TEM and imaged in the Vanderbilt Cell Imaging Shared Resource-Research Electron Microscopy facility as previously described. In brief, samples were fixed in 2.5% gluteraldehyde in 0.1M cacodylate buffer, pH7.4 at room temperature (RT) 1 hour then transferred to 4°C, overnight. The samples were washed in 0.1M cacodylate buffer, then incubated 1 hour in 1% osmium tetraoxide at RT then washed with 0.1M cacodylate buffer. Subsequently, the samples were dehydrated through a graded ethanol series and then 3 exchanges of 100% ethanol, followed by 2 exchanges of pure propylene oxide (PO). Samples were then infiltrated with 25% Epon 812 resin and 75% PO for 30 minutes at RT. Next, they were infiltrated with Epon 812 resin and PO [1:1] for 1 hour at RT then overnight at RT.

The samples are subsequently infiltrated with resin for 48 hours then allowed to polymerize at 60DC for 48 hours. 70-80nm ultra-thin sections were cut and collected on 300-mesh copper grids and post-stained with 2% uranyl acetate and then with Reynold’s lead citrate. Samples were subsequently imaged on the Philips/FEI Tecnai T12 electron microscope at various magnifications.

### Single Cell RNA sequencing

#### Tissue processing

Tissue sampling of the affected airway in the operating room in patients undergoing surgical treatment for their airway stenosis was preformed similar to tissue acquisition for 16S sequencing. After surgical exposure of the subglottis during direct laryngoscopy, the airway mucosa was washed with 10cc of sterile saline. After saline rinse, sterile upbiting cup forceps were used for mucosal biopsy. Without any break in sterility, a researcher received the mucosal explant in the operating room immediately after transfer from the surgical field. Tissue samples were placed in sterile room temperature saline and immediately transferred to the lab for tissue digestion. Biopsies were digested as previously described. In brief, samples were placed in an enzymatic cocktail [collagenase I/dispase II (1 μg/ml) tissue for 60 minutes followed by mechanical disruption. Tissue lysates were serially filtered 100- and 40-μm sterile filters (Fischer) and used as input for generation of scRNA-seq libraries.

#### scRNA-seq library preparation and next-generation sequencing

scRNA-seq libraries were generated using the 10x Chromium platform 3′ v2 or 5′ library preparation kits (10x Genomics) following the manufacturer’s recommendations and targeting 5000 to 10,000 cells per sample. Next-generation sequencing was performed on an Illumina NovaSeq 6000.

#### Reference Mapping, Cluster annotation, Differential Gene Expression (DEG), Gene Set Enrichment analysis (GESA)

10x Genomics Cell Ranger^13^ 5.0.0 was used to build reference genome index, map reads to reference genome (GRCh38-2020-A) and quantify genes. The cells labeled with sample-specific hashtags were demultiplexed by in-house scripts. scRNABatchQC^14^ was used for quality control. Cells were removed if they had unique features counts of less than 200 or larger than 8000, had fewer than 500 read counts and/or had greater than 40% mitochondrial read counts. Seurat^15^ was used for clustering analysis with sctransform based normalization. Cell type of each cluster was initially classified based on cell activity database^16^ and then manually refined based on cell type specific marker gene expression. edgeR^17^ was used to detect differential expression across conditions. Gene set enrichment analysis was performed using GSEA package^18^. Based on significantly differential expressed genes, Gene Ontology (GO) and pathway analysis was performed using WebGestaltR package^19^.

### Data and code accessibility

Raw and processed 16S sequencing data can be found under Bioproject. Raw and processed 10x Genomics data can be found on GEO using accession number GSE191128.

### In Vitro Cell Culture

Tissue biopsies acquired during operative endoscopy of 5 unique iSGS patients were used to create fresh single cell suspensions as described above. Suspensions were rested for 6 hours in RPMI Complete Subglottic Media [ RPMI + L-GLUT, 10% FCS + NEAA + IL-15 (25ng/ml) + 17B2 (1X10-12M Estrogen) ]. During the operative endoscopy 5mL of saliva was also collected from each matched iSGS patient and unrelated healthy control. These samples were used to generate an oropharyngeal microbiome based on published methods^20^. In brief, saliva samples were centrifuged at 2,000g for 5 mins to spin down large debris and eukaryotic cells. Next 1mL supernatant is removed from the sample and added to 1mL PBS (total of 2 mL) and stored anaerobically at 37degree C until use. After the period of rest, cells were cultured in the presence of a matched iSGS airway microbiome, the microbiome from an unrelated healthy subject, or left untreated. 24 hours after initiation stimulation, cells were washed twice in PBS and stained, and analyzed via flow cytometry.

#### Flow Cytometry

Single cell suspensions were prepared and adjusted to a concentration adjusted to 106/ml. To block nonspecific staining due to Fc receptor (FcR) binding, cells were preincubated with 5ul FcR Blocking Solution (Human TruStain FcX™, Biolegend inc, San Diego Ca. Cat. #422301) in 50ul PBS/2%FCS (Phosphate Buffered Solution, Fetal Calf Serum) per 1X10^6 cells for 10min at 4°C. The cells were not washed after FC blocking prior to the first staining step. Cells were then incubated 30 minutes at 4°C in a staining buffer (approx. 10^6 cells in 100μl of staining buffer). The staining buffer contained a pre-titrated, optimal concentration (≤ 5μg) of fluorescent monoclonal antibodies specific for a receptor or with an immunoglobulin (Ig) isotype-matched control respectively (see below for details). After the incubation, cells were washed 2x with 200ul of staining buffer and pelleted by centrifugation (250 X g for 5 min). Finally, cells were resuspended in PBS/2%FCS for flow cytometric analysis. All flow cytometry experiments were acquired with an LSR-II flow cytometer (BD Biosciences). Analysis was performed using FlowJo, LLC software (FlowJo, LLC, Ashland, OR).

Cells were gated by FSC/SSC, doublets were gated out, followed by exclusion of dead cells, and then cells were analyzed for characteristic surface protein expression. T cells were defined as CD45+ & CD326^neg^, CD3+, and either CD4+ or CD8+. In co-culture experiments CD154 expression was evaluated on CD4+ and CD8+ T cells.

Flow Cytometry Reagents Utilized

| Characteristic | Analyte | Detector | Reporter | Manufacture | Clone | Cat# |
| --- | --- | --- | --- | --- | --- | --- |
| Pan-Immune | CD45 | Anti- CD45 | AF700 | BioLegend | HI30 | 304024 |
| Pan Epithelial | CD326 (EPCAM) | Anti-CD326 | PE/Cy7 | BioLegend | 9C4 | 324222 |
| T Cells | CD3 | Anti-CD3 | BV605 | BioLegend | OKT3 | 317322 |
| CD4+ T Cells | CD4 | Anti- CD4 | PerCP | BioLegend | RPA-T4 | 300530 |
| CD8+ T Cells | CD8 | Anti-CD8a | FITC | BioLegend | RPA-T8 | 301050 |
| T-Cell Activation | CD154 | Anti-CD154 | APC/Cy7 | BioLegend | 24-31 | 310822 |
| Cell viability | DNA (compromised cells) | LIVE/DEAD™ | Fixable Aqua | Thermofisher | N/A | L34957 |

### Statistical Analyses

#### Microbiome / 16S sequencing

Our primary interest was comparison of the tissue microbiome of iSGS and post-intubation (iLTS) patients versus healthy controls. Additional secondary analyses tested the relationship of 1. disease severity (quantified as the number of procedures/year) and with microbial species in iSGS patients. To contextualize our findings with the published evidence in the field, we also compared our iSGS sequencing data with available published sequencing data for iSGS patients from airway mucosal swabs (from the Hillel et al. study)^11^ .

We set a read cutoff of 500 sequences per sample (2 standard deviations above the maximum number of reads retained in any of the negative controls). We calculated descriptive statistics and performed comparisons with non-parametric statistical tests using the R platform.^21^ Taxonomic descriptions, richness and alpha- diversity metrics were calculated with the R package vegan^22^ at the amplicon sequence variants (ASV) level. To control for differences in sequencing depths, counts were randomly rarefied to the lowest library size of all samples and then each microbial ecology index was computed. For each index, this rarefaction and computation process was repeated multiple times (n=400), and the results were averaged.

Beta diversity was assessed using the Bray-Curtis dissimilarity index with PermANOVA testing implemented in Adonis2 ^23^ to identify differences in community structure between iSGS cases, iLTS controls, and healthy samples at the ASV level. Principle coordinates analysis (PCoA) was performed in *vegan* in order to ordinate the dissimilarity data and plot it in two dimensional space. PCoA vectors and centroids were extracted with the *betadisper* function. Percent of variance explained by each PCoA axis was calculated by dividing its eigenvalue over the sum of all PCoA eigenvalues and multiplying by 100.

Secondary analyses testing associations between disease severity (procedures/year) with bacterial ASVs in iSGS were assessed using the R package DESeq2 (with p- or q-value as appropriate < 0.05).^24^ We eliminated ASVs that were detected on average <10 times, ASVs with a minimum quantile mean fraction <0.25, and ASVs with a minimum quantile incidence fraction <0.25. The absolute counts from the removed ASVs were aggregated into a category “other”, which was taken into account when computing simple proportions during data normalization but were otherwise discarded. This was done to reduce the penalty associated with multiple comparisons and to remove likely non-informative data.

The DESeq2 test uses shrinkage estimation for dispersions and fold changes to improve the stability and interpretability of estimates. This method models raw absolute counts of each taxon with a negative binomial distribution and uses the estimated depth of sequencing of each sample to scale the (unknown) relative abundance that is the parameter of the negative binomial distribution. Compared to using either simple proportion-based normalization or rarefaction to control for differences in sequencing depth, the DESeq2 test provides improved sensitivity and specificity.^25^ Default outlier detection and replacement was used as described.^24^ Reported q-values are the result of a Wald test with the Benjamini-Hochberg correction^26^ applied to adjust for multiple comparisons.

#### Additional Statistical Methods

Additional analyses were conducted with Prism (GraphPad Software). Nonparametric data were analyzed by Mann-Whitney U test for two variables and by Kruskal-Wallis test for greater than two variables, using Dunn’s post hoc analysis test. Categorical data were assessed by Fisher’s exact test. Correlations were assessed by linear regression. A P value less than 0.05 was considered statistically significant.

---

**Figure S1.**
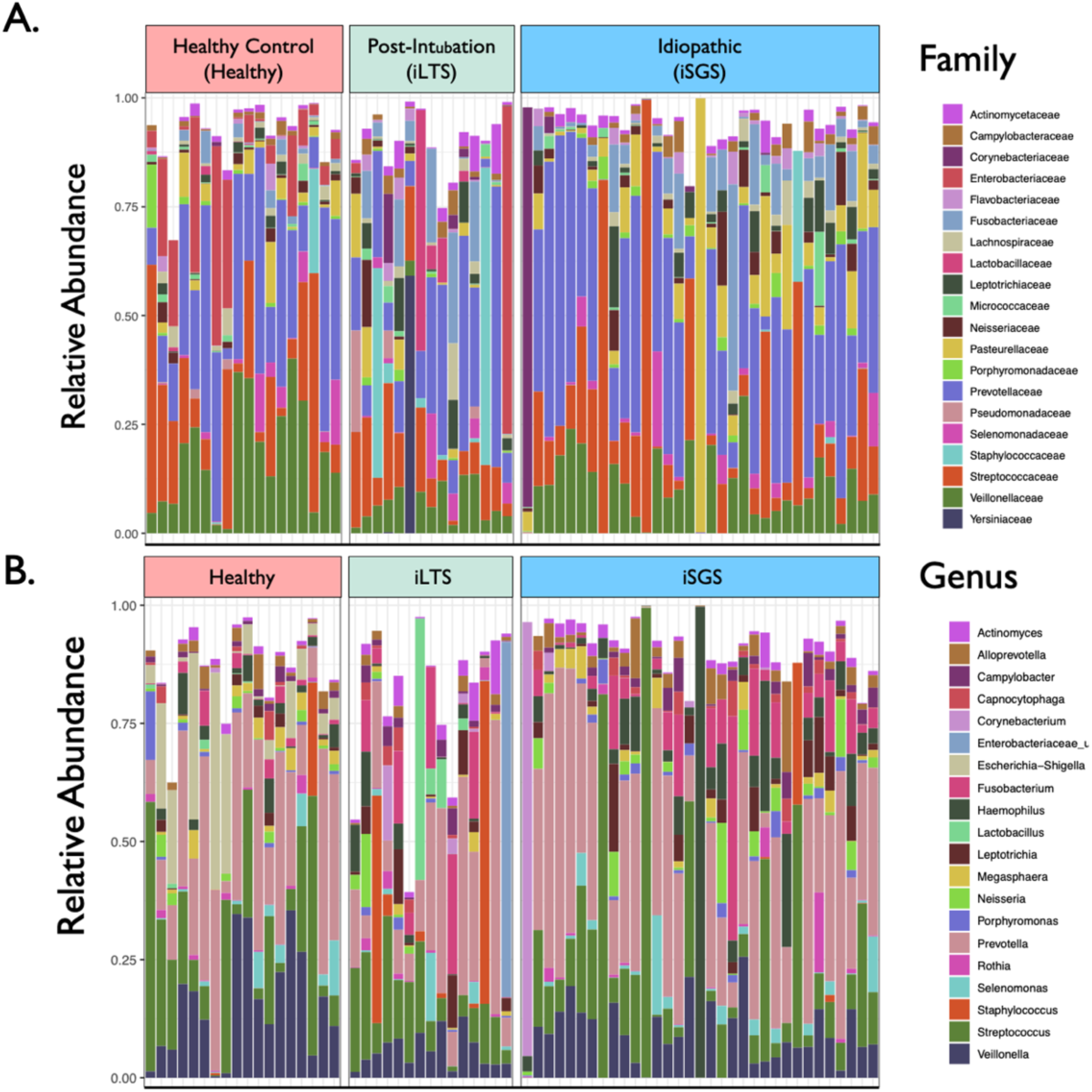
Relative bacterial abundance in iSGS cases and controls. Relative abundance of the top 20 most abundant A) families and B) genera in a stacked bar chart. Each sample represented as a unique column. Samples grouped into iSGS cases and post-intubation subglottic stenosis (iLTS) and healthy mucosal controls.

**Figure S2.**
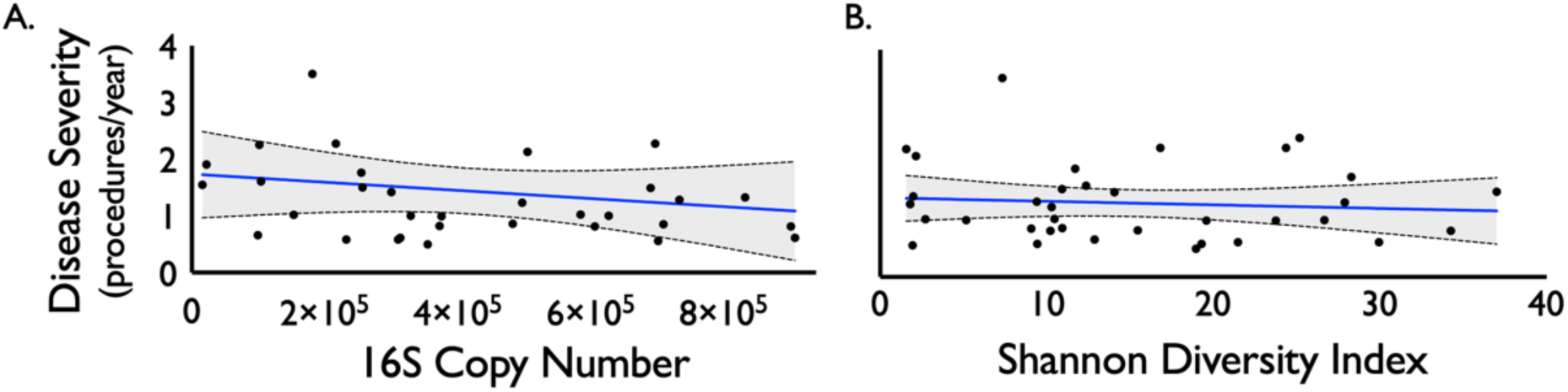
Relationship between iSGS disease severity and bacterial load and microbial diversity. There was not a significant association between iSGS disease severity (calculated as the number of procedures/year) and overall bacterial load (as quantified by 16S rRNA copy number, p-value = 0.953, Figure S2A). Nor was there an association between disease severity and alpha diversity (p-value = 0.453, Figure S2B) or richness (p-value = 0.6078, data not shown).

**Figure S3.**
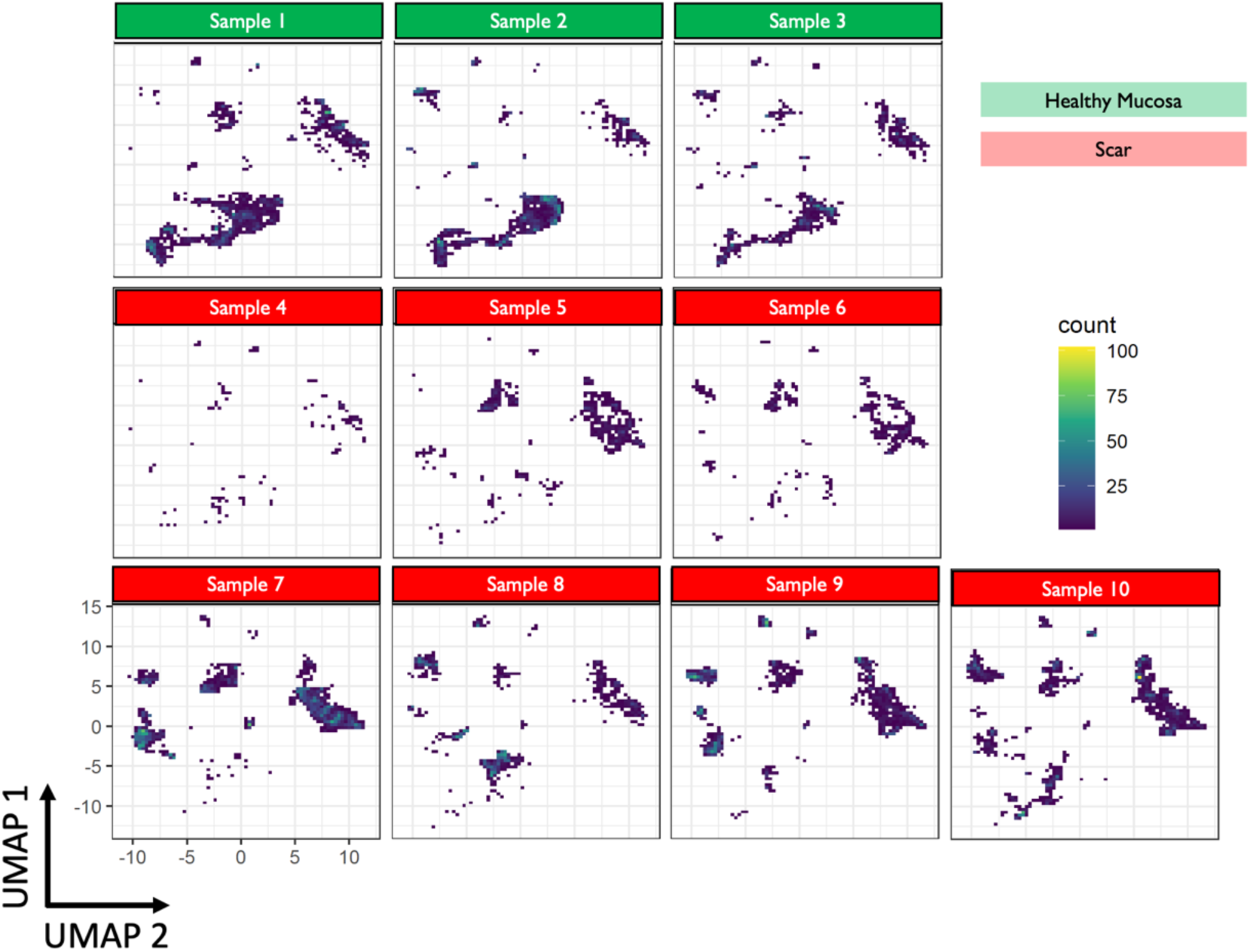
UMAP of individual samples. UMAP for each individual sample utilized (n=10: scar =7, matched healthy mucosa=3). There were not significant differences observe across biologic replicates.

**Figure S4.**
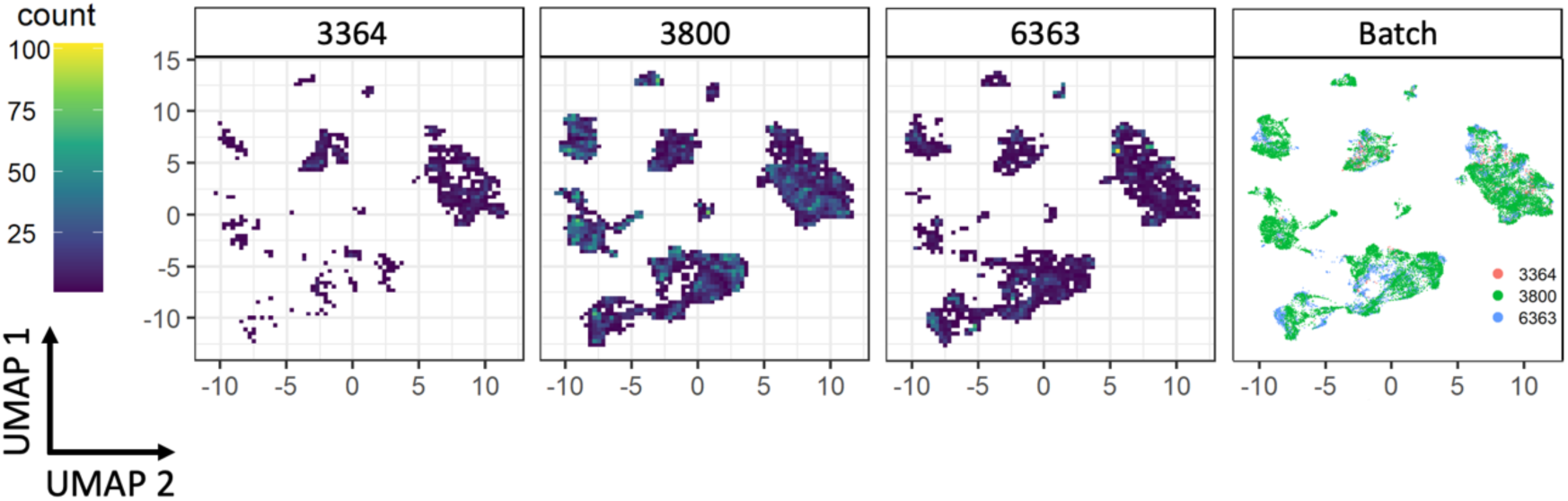
UMAP of sequencing batches. UMAP for each sequencing batch used (n=3). There were not significant differences observe across sequencing runs

**Figure S5.**
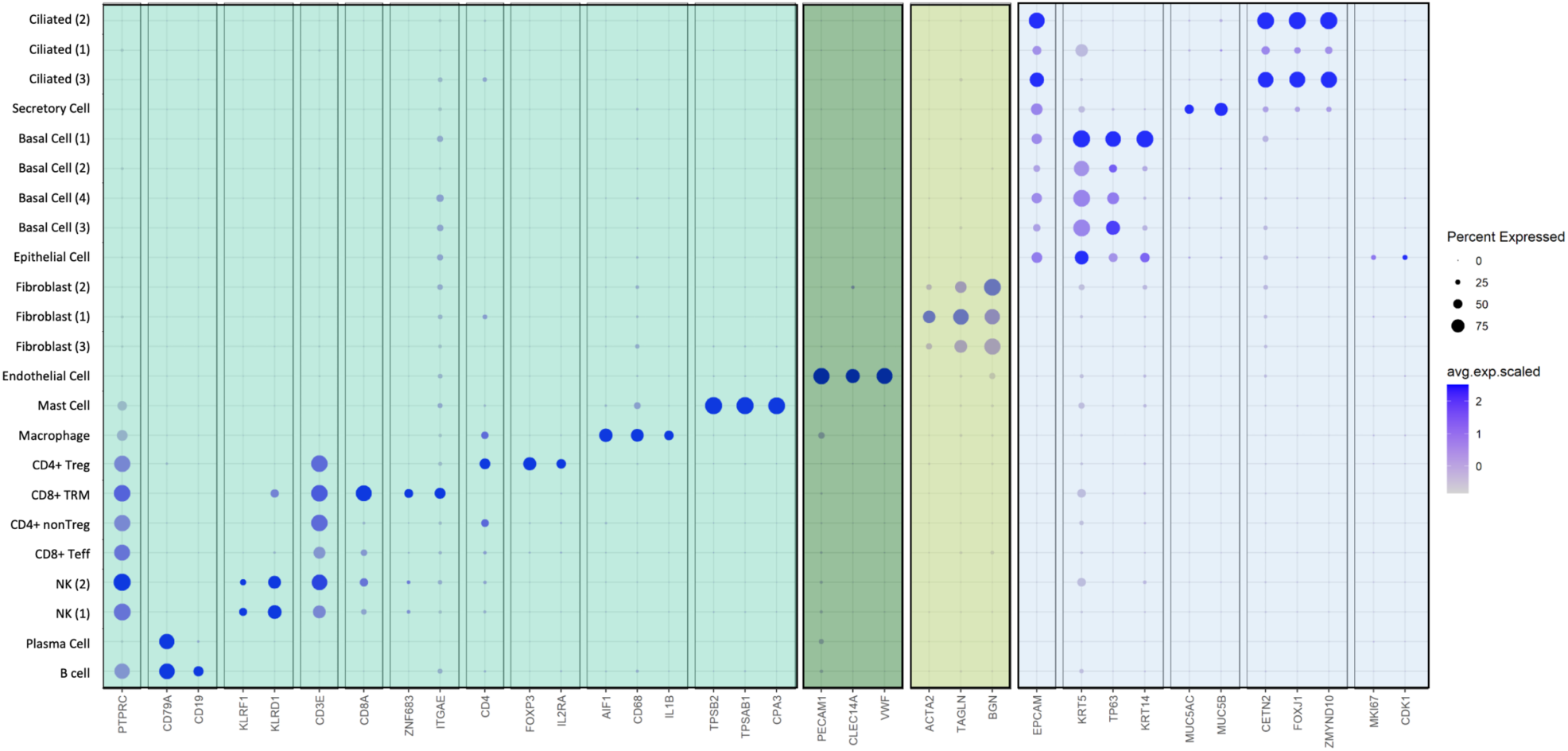
Expression of canonical cell-type markers among clusters. Canonical lineage specific marker expression confirming identify of proximal airway cell clusters. Size of dot indexed to percent expressed and color intensity reflects average scaled expression (deep blue depicting highest expression).

**Figure S6.**
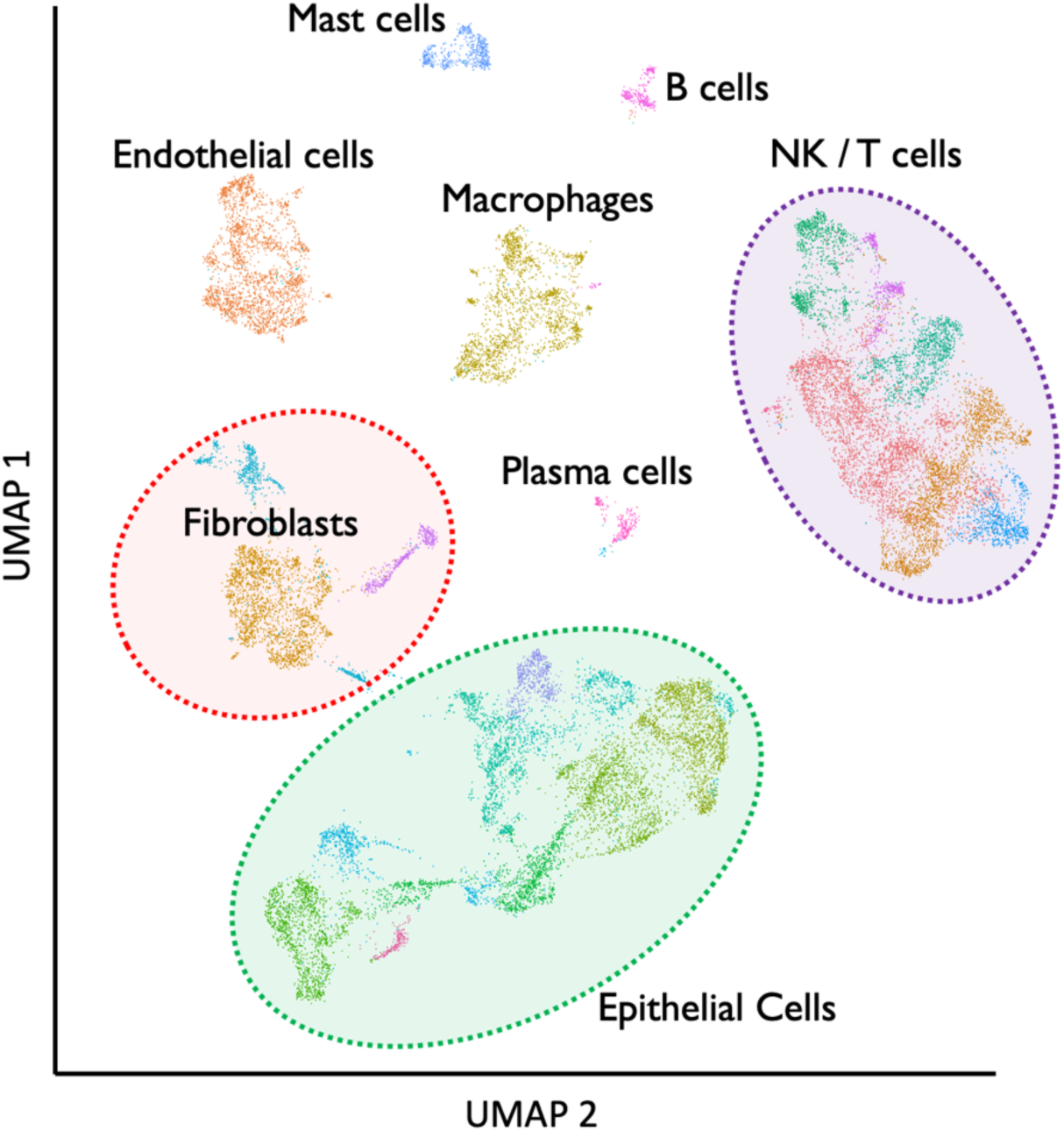
Global Cell Clusters. UMAP demonstrating cell types/states present in the proximal airway mucosal scar in iSGS patients. Labels derived from canonical marker expression (Figure S5).

**Figure S7:**
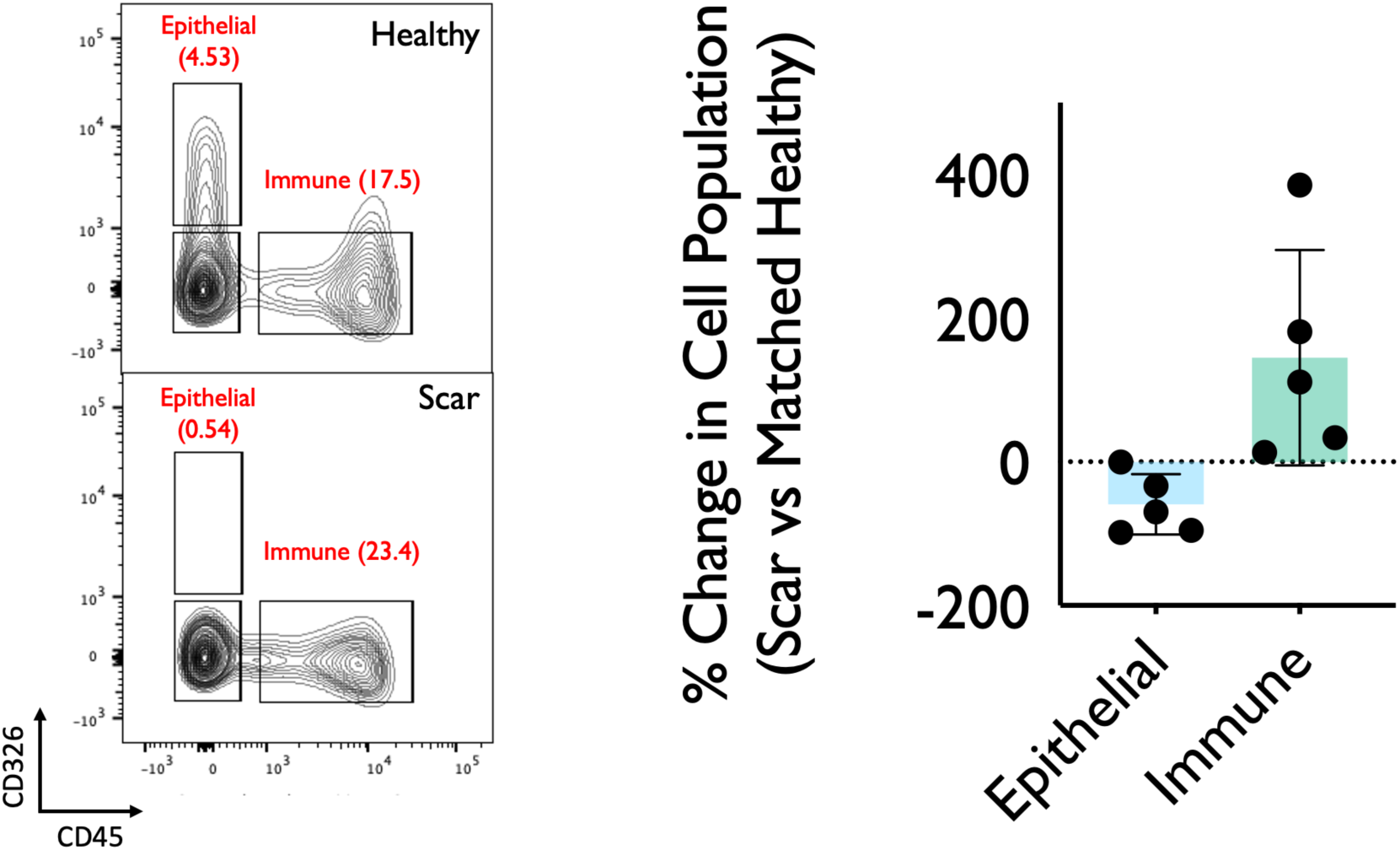
Flow Cytometry Confirming Epithelial Depletion and Immune Cell Increase in iSGS Airway Scar. Representative flow cytometry of fresh single cell suspension from matched iSGS airway scar and healthy mucosa (n=5) confirming depletion of epithelial cells and increase in immune cells within iSGS airway scar when compared to matched healthy control. Cells gated on FSC/SSC, singlets, live cells followed by CD45 (AlexaFlur® 647) and CD326 (EpCAM PE/Cy7). Graph depicting % change in cell population for matched scar/healthy mucosa for each of the five individual patients. Bar represents mean, error bars SEM, and dots show individual patients.

**Figure S8:**
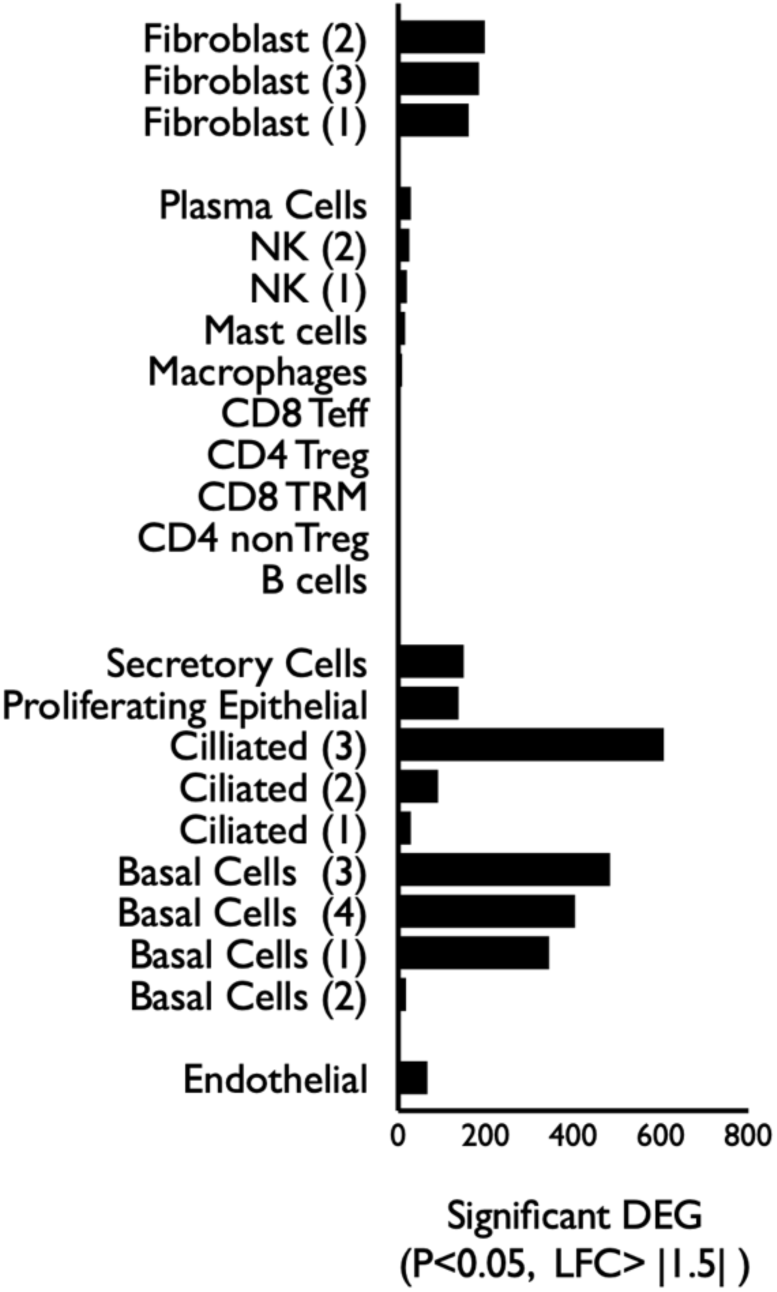
Differentially Expressed Genes (DEG) in Each Cell Cluster. Differentially expressed genes (DEG: P<0.05, log fold change > |1.5|) depicted for each cell type. Wide variability across the cell types with epithelial cell and fibroblast subsets showing the greatest number of DEG.

**Table S1.** Clinical Data for iSGS Cases and Controls Utilized in 16S Sequencing.

| Table S1. Patient Characteristics | iSGS<br>(n=50) | iLTS<br>(n=27) | Significance<br>(p) |  |
| --- | --- | --- | --- | --- |
| *Demographics* |  |  |  |  |
| Age (Mean years, 95% CI) | 47<br>(43 - 51) | 48<br>(41 - 54) | ns | <sup>1</sup> |
| Sex (% Female) | 100 | 66 | 0.001 | <sup>2</sup> |
| Race (% Caucasian) | 100 | 93 | 0.017 | <sup>2</sup> |
| Ethnicity (% Non-Hispanic or Latino) | 100 | 100 | ns | <sup>2</sup> |
| *Disease Morphology* |  |  |  |  |
| % Stenosis (Mean %, 95% CI) | 54<br>(48 - 60) | 73<br>(57 - 89) | 0.018 | <sup>1</sup> |
| Stenosis Length (Mean mm, 95% CI) | 12<br>(10 - 14) | 19<br>(15 - 22) | <0.001 | <sup>1</sup> |
| Distance below glottis (Mean mm, 95% CI) | 13<br>(12 - 15) | 21<br>(16 - 26) | <0.001 | <sup>1</sup> |
| *Treatment History* |  |  |  |  |
| Total Number of Surgical Procedures | 7.2<br>(5 - 9) | 7.7<br>(5 - 10) | ns | <sup>1</sup> |
| *Health / Endocrine Status* |  |  |  |  |
| Charlson Comorbidity Index (Mean, 95% CI) | 1.67<br>(0 - 0) | 2.71<br>(0 - 0) | 0.012 | <sup>1</sup> |
| GERD (% positive) | 46 | 48 | ns | <sup>2</sup> |
| DMII (% positive) | 12 | 62 | <0.001 | <sup>2</sup> |
| COPD (% positive) | 0 | 30 | <0.001 | <sup>2</sup> |
| *Follow-up Duration* |  |  |  |  |
| Years of Follow up (Mean months, 95% CI) | 71<br>(54 - 88) | 30<br>(14 - 46) | 0.002 | <sup>1</sup> |

Demographic and disease information for Idiopathic subglottic stenosis (iSGS), and post-intubation subglottic stenosis controls (iLTS). Mean (95% CI) for continuous variables. Covariates distinguishing diseases were sex (iSGS patients were all female) and ethnicity (iSGS patients were exclusively Caucasian). iLTS patients also had a longer length stenosis, with a greater degree of luminal obstruction, arising farther from the glottis. A greater percentage of iLTS patients had comorbid diabetes mellitus type II (DMII), and Chronic obstructive pulmonary disease (COPD). The median duration of patient follow-up was longer in iSGS patients. Tests used: 1: Kruskal-Wallis test; 2: Pearson test.

**Supplemental Table S2:** Demographics of Specimens Utilized for scRNAseq experiments.

| Patient ID | Sex | Diagnosis | Race |  |
| --- | --- | --- | --- | --- |
| 1 | Female | iSGS | Caucasian | *matched w/ 10* |
| 2 | Female | iSGS | Caucasian | *matched w/ 9* |
| 3 | Female | iSGS | Caucasian | *matched w/ 8* |
| 4 | Female | iSGS | Caucasian |  |
| 5 | Female | iSGS | Caucasian |  |
| 6 | Female | iSGS | Caucasian |  |
| 7 | Female | iSGS | Caucasian |  |
| 8 | Female | iSGS | Caucasian |  |
| 9 | Female | iSGS | Caucasian |  |
| 10 | Female | iSGS | Caucasian |  |

**Supplemental Table S3:**
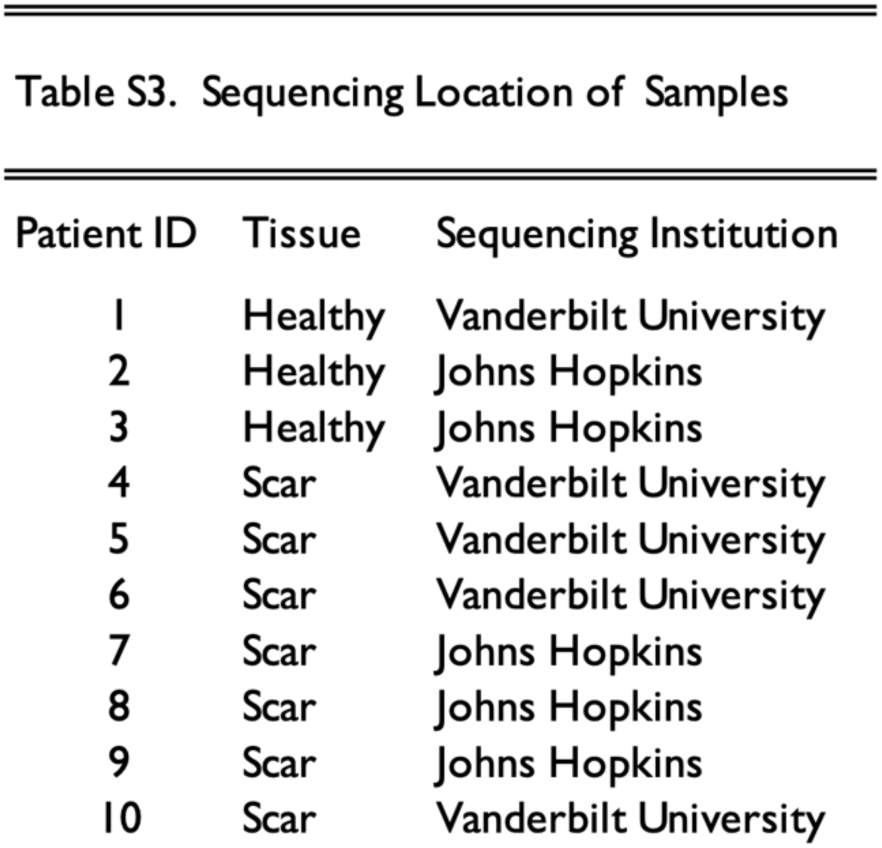
Sequencing Location of Samples utilized for scRNAseq experiments. Samples 1,4,5,6,10 were completed at Vanderbilt University and samples 2,3,7,8,9 were completed at Johns Hopkins. Both sites sequenced scar and matched healthy mucosal control

